# Polypore Mushroom Mycelia as an Adjunct to COVID-19 Vaccination: A Randomized Clinical Trial

**DOI:** 10.1101/2025.06.06.25327723

**Authors:** Gordon Saxe, Christine N. Smith, Shahrokh Golshan, Tatyana Shekhtman, Zolton J. Bair, Chase Beathard, Renee A Davis, Lauray MacElhern, Andrew Shubov, Daniel Slater, Lan K. Kao, Phoebe Senowitz, Stephen Wilson

## Abstract

Use of fungal mycelium as a vaccination adjunct may constitute a novel antiviral strategy to address newly emerging viruses. In a randomized, double-blind, placebo-controlled clinical trial, we evaluated safety and feasibility of fungal mycelium (*Fomitopsis officinalis* and *Trametes versicolor,* FoTv) as an adjunct to human COVID-19 vaccination, as well as its impact on vaccine side-effects and anti-SARS-CoV-2 antibodies (Abs). We evaluated safety, feasibility, vaccine side-effects (across 5 days), and anti-SARS-CoV-2 Ab levels (across 6 months). Safety metrics were similar for the FoTv (N=52) and Placebo (N=38) groups and the approach was feasible. Participants with detectable Abs (from prior COVID antigen exposure) were classified as “COVID-Exposed” and those with undetectable Abs as “COVID-Naive.” FoTv, versus Placebo, significantly reduced side-effects in COVID-Naive, but not in COVID-Exposed individuals. In the COVID-Naive FoTv group, Ab responses were preserved across 6 months, an effect not observed among other groups. Adjunctive FoTv was safe, feasible, and reduced vaccine side-effects without compromising (and possibly increasing) Ab levels up to 6 months in participants without previous SARS-CoV-2 exposure. Use of fungal mycelia was successfully tested as an approach to prevent a novel pandemic virus (SARS-CoV-2), with potential application to H5N1/Bird Flu and other emerging viruses.

## INTRODUCTION

### Lessons from the COVID-19 pandemic

As we take stock of the societal response to COVID-19 and plan for future pandemics, it has become clear that the medical community needs to identify successful strategies to minimize morbidity, mortality, and global disruption that could result both from newly evolving strains of SARS-CoV-2 as well as from other viruses or diseases (e.g., bird flu, monkey pox, Zika virus, Hantavirus, dengue fever, etc.) that could broadly threaten human health. Various safe and effective strategies, encompassing primary prevention, vaccination, antiviral medications, and diet/lifestyle modifications, are urgently needed to protect against existing and emerging strains of infectious diseases.

Strategies that incorporate vaccines/antiviral medication as the first lines of defense will likely be relied upon. Although vaccination has a long history of successful use in prevention of many viral and other illnesses, concerns have been raised about durability and cross-protective immunity conferred by mRNA and other forms of vaccination against SARS-CoV-2, as well as about vaccine-associated side effects. Unfortunately, antiviral medications also cannot be relied upon as a primary strategy because they: (a) may be in short supply/inaccessible to many individuals (for financial reasons or due to problems with distribution); (b) may encounter drug resistance because of overuse (e.g., prophylactic administration to livestock); and (c) are only likely to be effective if taken during a limited time window during the disease course (shortly after exposure or, at most, immediately after symptom onset).

In addition to COVID-19, other emerging infectious diseases also warrant close attention. Among these, highly-pathogenic avian influenza A (HPAI) is of particular and growing concern. HPAI, caused by a viral subtype of H5N1, has led to a serious form of bird flu with symptoms in humans ranging from mild (e.g., conjunctivitis, flu-like symptoms) to severe/fatal (e.g., respiratory failure). Between 2003 and 2024, 463/896 cases of HPAI due to H5N1 were fatal (52% case-fatality rate per reports from 24 countries).^1^ According to the Centers for Disease Control and Prevention (CDC), a new H5N1 clade (2.3.4.4b) was first detected in animals in 2021, with the first human case reported in January 2022. Although these cases have generally resulted in mild illness, this clade has been detected in over 500 avian and mammalian species globally and, more recently, in poultry, cattle, swine, and dairy products. The increasing cross-species transmissibility (avian-mammal, mammal-mammal, avian-human, and mammal-human) and broadening host range of this clade raise concern that sustained human-human transmission of this virus could soon develop^2^. This, in tandem with continuing outbreaks of this clade in animals and the high case-fatality rate of previous clades, has led to worry that a global pandemic virus more lethal and disruptive than SARS-CoV-2 could emerge.

### Vaccination as a Strategy to Address Newly Emerging or Potentially Pandemic Infectious Diseases

Vaccination was epidemiologically impactful in resolving the COVID-19 pandemic. This was possible because the virus was identified early and new technology (i.e., mRNA vaccines) was quickly and specifically designed to exploit points of viral vulnerability (e.g., Spike protein). Yet, there were shortcomings to this strategy. First, concerns persisted regarding side effect frequency and severity. Second, vaccine efficacy was often suboptimal, particularly in immunocompromised individuals, and tended to wane over time. Third, new strains of SARS-CoV-2 continued to emerge, resulting in further reductions in vaccine efficacy and durability, and need for more frequent boosters.

Not surprisingly, a significant portion of the population expressed vaccine hesitancy. According to a CDC report on a December 2020 survey coinciding with initial rollout of COVID-19 vaccines, approximately 30% of people not intending to be vaccinated identified “concern about side effects and safety” as their primary reason^3^. Later, a similar proportion identified concern about vaccine side effects as their reason for not receiving a booster^4^. Vaccine-related side effects are commonly mild and short-term, but are occasionally serious or longer-term^5^. Frequency of side effects, particularly common short-term ones, may be related to inflammation and heightened immunoreactivity to vaccine adjuvants^6^ and may worsen with re-exposure^7,8^. Because reactogenic side effects impede public health goals of widespread vaccination, an important challenge is to identify adjunctive agents/approaches that reduce excessive reactogenicity without compromising immunogenicity.

Vaccine immunogenicity is influenced by various factors related to immunological status, including diabetes,^9^ chronic renal failure,^10^ chronic liver disease,^10,11^ steroid use,^12^ prior infection,^13^ age,^14,15^ gender,^16^ nutritional status,^10,17^ obesity,^18^ gut microbial balance,^19^ antibiotic use,^20^ and prior vaccination/infection.^21^ Post-vaccine levels of SARS-CoV-2 anti-Spike protein antibodies (Abs) are correlated with and may serve as surrogates for protection against infection.^22^ Unfortunately, levels of neutralizing Abs are insufficient to afford robust protection to many immunocompromised individuals.^23,24^ Further, Ab levels wane over time, marked by increasing rates of COVID-19 infection within 2-6 months post-vaccination.^25,26^

During the pandemic, extensive use of vaccines to prevent infection by SARS-CoV-2 (a virus to which the population was initially immunologically naive) revealed widespread vaccine hesitancy (particularly related to side effects concerns), and safety and efficacy concerns. Such concerns need to be addressed because vaccines will likely serve as a first defense to address new SARS-CoV-2 variants or other emerging viruses with pandemic potential. The use of fungal mycelium, in conjunction with immunization, may offer a novel strategy to reduce adverse side effects without compromising (or perhaps even enhancing) immunogenicity, thereby enhancing vaccine acceptance and efficacy.

### Vaccine Enhancement with Agaricomycete Fungi

Agaricomycete fungi, consisting of mycelium (root-like network) and fruit bodies, have been studied as vaccine adjuvants in animal models and used in veterinary care; examples include *Phellinus linteus* (Pl)^27^ mycelium extract, turkey tail *(Trametes versicolor,* Tv) polysaccharides including polysaccharide-K (PSK),^28^ and *Pleurotus ostreatus* lectin.^29^ Mice receiving intranasal coadministration of influenza A vaccine and an adjuvant of Pl mycelium extract and then challenged with variant H5N1 viruses, showed significantly reduced viral titers and greater survival rates compared to non-adjuvanted controls^27^. More recently, coadministration of SARS-CoV-2 vaccine (BNT162b2; Pfizer-BioNTech) and an adjuvant Tv mycelial extract (in combination with an extract of the Chinese herb *Astragalus membranaceus)* in mice, improved antigen (Ag)-binding efficacy against Spike proteins of SARS-CoV-2.^30^ In both studies, mycelial adjuvants to vaccination not only demonstrated efficacy against the targeted virus, but also induced cross-protection against variant strains of each respective virus. These findings suggest that agaricomycete fungal preparations may have appeal as candidate vaccine adjuvants/adjuncts.

Tv, a polypore fungus, has also demonstrated immunomodulatory,^31,32^ anti-cancer,^33–35^ and antiviral activities.^36,37^ Tv mycelium modified in-vitro expression of human monocytes and lymphocytes as well as pro-inflammatory, anti-inflammatory, and antiviral cytokines.^31^ Tv has been investigated in at least thirteen clinical trials for cancer, many focusing on mycelium-derived PSK, and demonstrating potential efficacy across various cancer types including breast, colorectal, gastric, and esophageal.^35^ Moreover, in breast cancer patients undergoing post-chemotherapy radiation, Tv increased lymphocyte and CD8+ T-cell counts, as well as tumoricidal activity of natural killer cells.^34^ Tv has consistently demonstrated anti-influenza activity, and in a comparative study of multiple mushroom species, Tv mycelium emerged as the most promising candidate for anti-influenza and anti-herpetic applications, owing to its potent therapeutic efficacy and low toxicity.^38,39^ Additionally, Tv mycelium was shown to reduce viral titers in a clinical trial of honeybees, supporting immunity by conferring a 75-fold reduction in Lake Sinai virus (LSV) in vivo.^37^

Another polypore, agarikon (*Fomitopsis officinalis* syn. *Laricifomes officinalis*, Fo), has been evaluated for the medicinal potential^40^ of both its mycelium and fruit bodies, demonstrating wide-ranging antiviral, antibacterial, anti-cancer, and anti-inflammatory effects^41^ suggestive of immunological activity.^42^ Fo exhibited promising anti-cancer activity in zebrafish, activating the immune system to inhibit angiogenesis through toll-like receptor and vascular endothelial growth factor signaling pathways.^43,44^ Extracts of Fo mycelium neutralized influenza A viruses including H5N1 in vitro, reducing infective virus titers at concentrations that were not cytotoxic^45^. In addition, Fo demonstrated in vitro activity against HSV, H5N1, H3N2, and Orthopox viruses,^45–48^ and bacteria including *Staphylococcus aureus* and *Mycobacterium tuberculosis*.^47,49^ Further, Fo triterpenoids exhibited anti-inflammatory effects in RAW-264.7 murine macrophages.^50^

Various polypore fungi, including Tv and Fo, demonstrate immunomodulatory effects, possibly through increased production of IL-1 receptor antagonist (IL-1Ra),^31^ a known immunoregulatory mediator. Normally, differentiation of B-cells to memory phenotypes occurs after IL-1 levels have reached their post-vaccination peak. Fungi, by increasing IL-1Ra, may enhance this differentiation process. This process may occur directly (through inhibition of IL-1 signaling via IL-1Ra), and indirectly (through attenuation of IL-6 and TNF-α, thereby supporting germinal center function and differentiation of T follicular helper cells). Notably, Fo and Tv, evaluated in the current study in combination (FoTv) as an adjunct to COVID-19 vaccination, has also undergone examination as a treatment for active COVID-19 infection in a randomized, placebo-controlled clinical trial of patients with mild-to-moderate illness.^51,52^

The primary objective of the current study was to examine effects of adjunctive short-term oral supplementation with FoTv versus Placebo to standard COVID-19 vaccination on vaccine reactogenicity and immunogenicity. We specifically hypothesized that:

H1: Post-vaccination FoTv treatment would be (a) safe and (b) feasible.

Because we anticipated that FoTv’s impact on vaccine side effects and Ab levels would be most readily detected among participants without prior SARS-CoV-2 Ag exposure (from prior infection or vaccination), outcomes in participants with or without exposure were examined separately. This approach reflects the findings that (1) prior exposure to Ag from either SARS-CoV-2 infection or COVID-19 vaccination influences temporal dynamics of primary and memory Ab responses^21^ and (2) repeated exposure to vaccine adjuvant increases severity of vaccine side effects.^7,8^ Therefore, we hypothesized that:

H2: Individuals without prior SARS-CoV-2 Ag exposure who received FoTv would (a) exhibit the largest reductions in vaccine-related side effects relative to all other treatment/exposure subgroups, and (b) would experience this reduction without adversely impacting their serum Ab levels.

In summary, the overall purpose of this study was to test, in the context of vaccination during the COVID-19 pandemic, the concept that fungal mycelium could serve as a safe, feasible, and effective vaccine adjunct. If so, it might serve as an effective tool for preventing infection from new strains of COVID-19 or in mitigating future pandemics from other emerging infectious diseases.

## RESULTS

Participants were enrolled between June 2021-January 2022, with the last follow-up visit in June 2022. Ninety participants were enrolled, 52 receiving FoTv and 38 receiving Placebo (Figure 1). Mean participant age was 39.0±15.6 years. Participants were: 57.3% female; 74.2% Caucasian, 19.1% Asian, 3.4% African American, 3.4% other race; and 22.5% Hispanic. Because of 2:1 FoTv-to-Placebo randomization ratio early in the study, together with vaccine policies then in place prioritizing older individuals, the FoTv group had a larger proportion of older individuals. Beyond age, there were no significant differences between FoTv and Placebo groups in demographic/clinical characteristics (Table 1) or efficacy variables (side effects or Abs) at baseline (all p-values >0.206). Accordingly, age was included as a covariate for analyses of continuous variables. Seventeen participants (19%) did not complete all study visits: Between days 14-28/42, one participant relocated, and another was lost to follow-up. Between days 28/42-6 months, 15 additional participants were lost to follow-up. Dropout was not significantly associated with Treatment Group (χ^2^ =0.201, df=1, p=0.654) and baseline efficacy measures were not significantly different between completers and non-completers (p-values >0.592). All participants were included in analyses regardless of if/when they dropped out.

**Figure 1.**
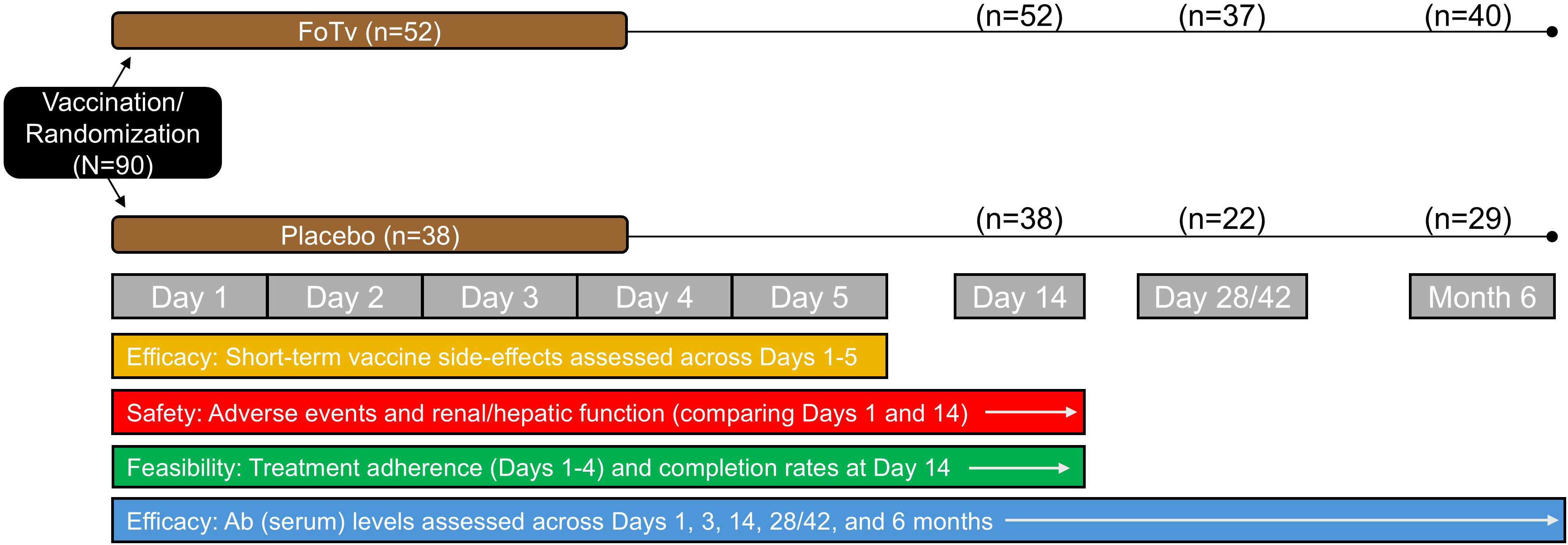
Design of Clinical Trial. At time of COVID-19 vaccination, participants were randomized to receive either a 4-day regimen of FoTv (*Fomitopsis officinalis/Trametes versicolor*) or Placebo on Day 1 (baseline). Vaccine side effects were assessed across Days 1-5. Adverse events were assessed across Days 1-14 and renal/hepatic function were assessed on Days 1 and 14. Feasibility analyses examined treatment adherence (Days 1-4) and completion rate at Day 14. Antibody (Ab) levels were assessed on Days 1, 3, 14, and 28/42, and at 6 months. Note that Day 3 reflects a time point 3 days after a booster vaccine, Day 28/42 reflects a time point 28 days after vaccination for the one-dose vaccine, or 42 days after the second dose of the two-dose vaccine.

**Table 1.** Patient Demographics and Baseline Clinical Characteristics.

| Characteristic | FoTv (n=52) | Placebo (n=38) | Statistic | df | p value |
| --- | --- | --- | --- | --- | --- |
| | Mean/Count<br>(SD/%) | Mean/Count<br>(SD/%) | F or $\chi^2$ | | |
| Age | 43.1 (15.6) | 33.5 (13.9) | 8.98 | 1,88 | 0.004 |
| Body Mass Index | 27.4 (7.8) | 25.3 (4.8) | 2.20 | 1,87 | 0.142 |
| Sex |  |  | 0.17 | 1 | 0.680 |
| Female | 31 (59.6%) | 21 (55.3%) |  |  |  |
| Male | 21 (40.4%) | 17 (44.7%) |  |  |  |
| Self-reported race |  |  | 4.69 | 3 | 0.196 |
| Asian | 9 (17.3%) | 8 (21.1%) |  |  |  |
| Caucasian | 41 (78.8%) | 26 (68.4%) |  |  |  |
| African | 2 (3.8%) | 1 (2.6%) |  |  |  |
| Other | 0 (0%) | 3 (7.8%) |  |  |  |
| Self-reported ethnicity |  |  | 0.327 | 1 | 0.567 |
| Hispanic | 11 (21.2%) | 10 (26.3%) |  |  |  |
| Not Hispanic | 41 (78.8%) | 28 (73.7%) |  |  |  |
| Vaccine Type |  |  | 1.47 | 2 | 0.480 |
| Moderna | 14 (26.9%) | 13 (34.2%) |  |  |  |
| Pfizer | 28 (53.8%) | 21 (55.3%) |  |  |  |
| Johnson & Johnson | 10 (19.2%) | 4 (10.5%) |  |  |  |
| Renal Function (eGFR) | 74.5% | 92.1% | 4.57 | 1 | 0.032 |
| Hepatic Function |  |  |  |  |  |
| AST | 96.2% | 100.0% | 1.50 | 1 | 0.507* |
| ALT | 82.7% | 89.5% | 0.82 | 1 | 0.366 |
| ALP | 100.0% | 94.7% | 2.78 | 1 | 0.176* |

*Note.* F-values and Chi-square statistics are reported for continuous and categorical variables, respectively. \*Due to the small sample size for cells in these analyses, probability values for Fischer's Exact tests are reported in lieu of Chi-square tests. AST=Aspartate Aminotransferase, ALT=Alanine Transaminase, ALP=Alkaline Phosphatase, and eGFR=adjusted glomerular filtration rate.

### Safety

No adverse events were reported in either FoTv or Placebo group. At baseline (and day 14, χ^2^=4.63, p=0.032), only renal function (percent of participants with normal eGFR) was significantly higher in the Placebo group. However, this inherent baseline characteristic of participants randomized to FoTv (vs. Placebo) possibly reflected their older mean age (Table 1). Importantly, there were overlapping CIs for the percentage of participants in each Treatment Group transitioning from normal to abnormal renal or hepatic function across this timeframe (Table 2).

**Table 2.** Percentage of Participants Transitioning from Normal Renal or Hepatic Function at Baseline to Abnormal at Day 14.

| Day 1 versus Day 14: |  |  |  |  |
| --- | --- | --- | --- | --- |
| Percentage Transitioning from Normal to Abnormal Function |  |  |  |  |
|  | FoTv |  | Placebo |  |
|  | % Transitioning | 95% CI | % Transitioning | 95% CI |
| Renal Function |  |  |  |  |
| Adjusted eGFR | 10.5 | 2.9-24.8 | 2.9 | 0.1-14.9 |
| Hepatic Function |  |  |  |  |
| AST | 2.0 | 0.1-10.6 | 2.6 | 0.1-13.8 |
| ALT | 4.7 | 0.6-16.2 | 5.9 | 0.7-19.7 |
| ALP | 1.9 | 0.0-10.4 | 0.0 | 0.0-10.6 |

*Note.* The Normal to Abnormal transition analysis reflects the percentage of participants with normal function at Day 1 that transitioned to abnormal at Day 14 compared to participants with normal function at both Days, across the Treatment Groups. AST=Aspartate Aminotransferase, ALT=Alanine Transaminase, ALP=Alkaline Phosphatase, and eGFR=glomerular filtration rate.

### Feasibility

Feasibility analyses examined eligibility for 174 participants, of whom 84 were excluded, and 90 were enrolled (Figure 2). After enrollment, all participants (100%) completed days 1-14 of the study (Figure 2). Treatment Adherence was high (96.0%±7.3%, CI: 94.4-97.5%); and was similar for FoTv (96.4%±6.7%, CI: 94.5-98.3%) and Placebo (95.4%±8.1%, CI: 92.7-98.1%).

**Figure 2.**
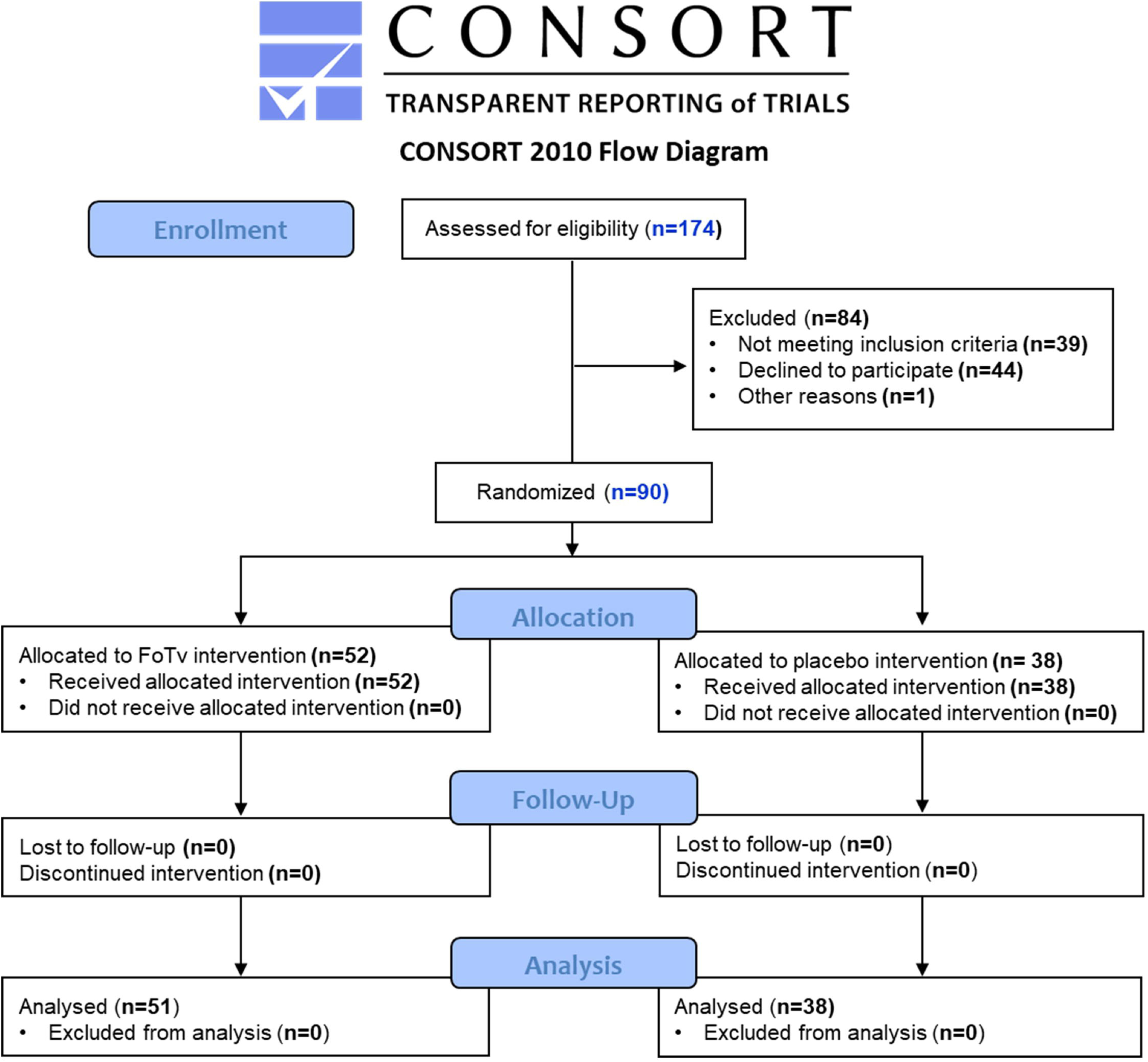
Consort Diagram for Clinical Trial Safety and Feasibility Data from Day 1 (baseline) to Day 14.

### Efficacy

Prior to efficacy analyses, five participants (2 Placebo, 3 FoTv) who had ≥1 extreme outlier Ab level (>3 SD above or below mean) were excluded, resulting in the following groups: FoTv: COVID-Naive (n=19), COVID-Exposed (n=30); Placebo: COVID-Naive (n=11), COVID-Exposed (n=25). Efficacy analyses examined Treatment Group by COVID-Exposure Status by Day interactions. Follow-up LME analyses were carried out separately for COVID-Exposure Status strata and Treatment Groups.

#### Side Effects

Presentation of vaccine-related side effects (over the first 5 days post-vaccination) depended on both prior COVID exposure and FoTv versus Placebo assignment. There was a strong trend for a Treatment Group by COVID Exposure Status by Day interaction (Days 1-5) for Side effect Count (F_(8,325)_=1.898, p=0.060; Figure 3A), but not for Side effect Severity (F_(8,309)_=1.156, p=0.326). Within COVID Exposure Status strata, there was a significant Treatment Group by Day interaction for COVID-Naive (F_(4,112)_=2.956, p=0.023), but not COVID-Exposed (F_(4,212)_=0.436, p=0.782), stratum for Side effect Count. Follow-up comparisons indicated COVID-Naive FoTv group reported significantly fewer side effects than Placebo on days 3 and 5 (F-values_(1,28)_ > 6.527, p-values < 0.017; Figure 3A). Within Treatment Groups, there was no significant COVID Exposure Status by Day interaction for FoTv groups (F_(4,188)_=0.861, p=0.488). However, there was a significant interaction for Placebo groups (F_(4,136)_=2.692, p=0.034).

**Figure 3.**
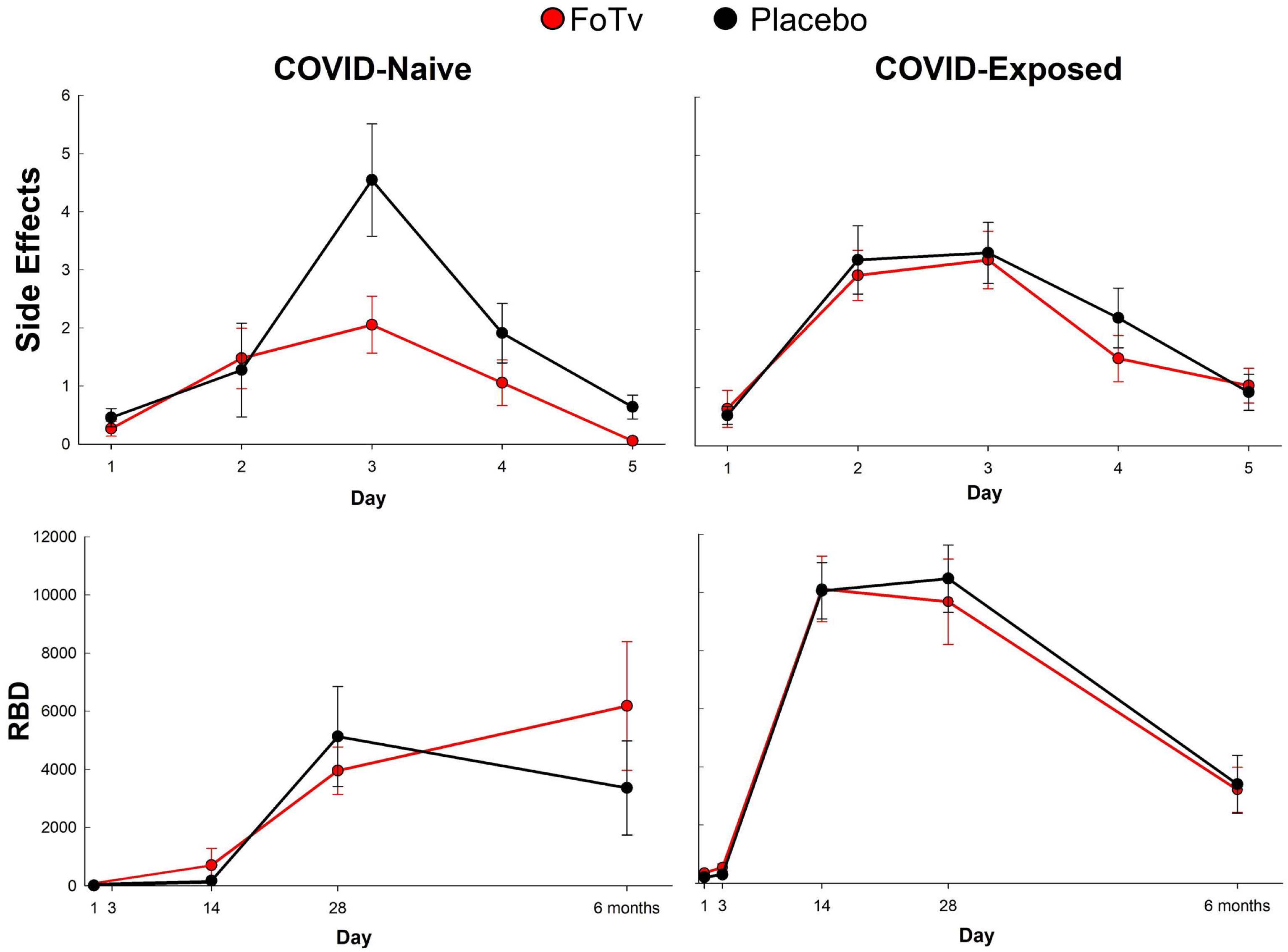
Efficacy results for Side Effects and Antibodies. **A.** Side Effect Counts across days 1 to 5. FoTv (red) and Placebo (black) groups for participants who were COVID-Naive (left) or COVID-Exposed (right) prior to vaccine/booster. The COVID-Naive FoTv group (n=19) exhibited a significantly lower number of side effects across days 1-5 relative to the Placebo group (n=11), whereas the COVID-Exposed FoTv (n=30) and Placebo (n=25) groups did not. **B.** RBD antibody response for groups as described in panel A. The COVID-Naive FoTv group exhibited significant increases from day 14 to 6 months, whereas the COVID-Exposed FoTv group exhibited significant decreases across these days (red lines). By contrast, the Placebo groups (black lines) did not significantly differ from each other. Error bars reflect SEM. RBD (anti-SARS-CoV-2 receptor binding domain) units are BAU (binding antibody units)/mL.

#### Antibodies

Ab levels (across 6 months) also depended on both prior COVID exposure and FoTv vs. Placebo assignment. Four participants who either contracted COVID-19 or received a COVID-19 booster before the 6-month timepoint were excluded from the 6-month efficacy analyses. There was a significant Treatment Group by COVID Exposure Status by Day (3, 14, 28/42, and 6 months) interaction for receptor-binding domain (RBD; (F_(2,150)_=6.374, p=0.002; Figure 3B) and Spike (F_(2,157)_=4.286, p=0.002) Abs. Results remained significant when outliers were included. Within each COVID Exposure Status stratum, there were no significant Treatment Group by Day interactions (all F-values_(1,53-144)_<1.080, all p-values >0.303). Within Treatment Groups, there were significant COVID Exposure Status by Day interactions for FoTv (RBD: F_(1,87)_=12.612, p<0.001; Spike: F_(1,91)_=8.358, p=0.005), but not Placebo (RBD: F_(1,65)_=1.379, p=0.245; Spike: F_(1,65)_=1.170, p=0.283). Follow-up, pairwise comparisons of Ab levels across Days indicated the COVID-Naive FoTv group exhibited significant RBD increases across timepoints (Day 14-28/42; 28/42-6 months; all t >2.314, df=31-33, all p<0.027), whereas the COVID-Exposed FoTv group (Day 14-6 months; t=-8.561, df=39, p<0.001) exhibited significant decreases. A pattern similar to RBD was observed for Spike Ab. In the COVID-Naive FoTv group there was a significant increase across timepoints (Day 14-28/42; 28/42-6 months; all t-values>3.527, df=31-32, all p-values<0.001). However, there were significant decreases for the COVID-Exposed FoTv group (Day 14-6 months; (t=-11.996, df=39, p<0.001). The strength of the FoTv effect on Ab levels was even stronger when five outlier participants with extremely high Ab levels were included in the analysis.

Ab response durability analyses explored changes from the Placebo group peak response (Day 28/42) to end-of-study (6 months). The COVID-Naive FoTv group exhibited increasing Ab levels, whereas other groups exhibited decreasing Ab levels (Figure 3B, Supplemental Table 3). There were trends toward more durable treatment-related Ab responses in the COVID-Naive stratum (Treatment Group by Day interaction RBD: F_(1,25)_=2.706, p=0.113; Spike: F_(1,24)_=2.829, p=0.105). However, no trends were observed in the COVID-Exposed stratum (RBD: F_(1,35)_=0.078, p=0.782; Spike: F_(1,36)_=1.206, p=0.279).

## DISCUSSION

This study addressed whether FoTv could serve as a safe, feasible, and effective adjunct to COVID-19 vaccination. Short-term (4-day) oral FoTv administration was safe (Table 2) and feasible (Figure 2). In those who had not been exposed to COVID-19 infection or received prior vaccination (i.e., COVID-Naive individuals), FoTv also reduced short-term (5-day period post-vaccination) side effects (Figure 3A) while preventing the normal decay of vaccine Ab responses at 6 months (end of study; Figure 3B). If anything, FoTv may have *increased* Ab response durability, in that Placebo Ab levels declined (after peaking at 28/42 days) while FoTv Ab levels continued to increase up to 6-months in COVID-Naive individuals. FoTv constituted a safe, acceptable adjunct to vaccination for COVID-19 and, an optimal characteristic vaccine adjuvant or adjunct, it may have decreased reactogenicity while preserving or increasing immunogenicity.

This pattern, minimized short-term side effects coupled with increased Ab durability, suggests immunomodulation, and is consistent with what is known about the immunobiology of fungi. Despite earlier speculation that fungal polysaccharides might stimulate excessive reactogenicity and potentially trigger cytokine storm during acute COVID-19 infection,^53^ a recent clinical study of COVID-19 patients instead found fungal β-glucan treatment decreased inflammatory markers (IL-6 and d-Dimer).^54^ Further, this idea is reinforced by the fact that polypore fungi contain various bioactive peptides,^55^ compounds known to regulate cytokine responses required for an optimal adaptive immune response.^56^

In COVID-Naive stratum, FoTv produced more gradual and ultimately higher peak Ab responses than Placebo (Figure 3B), suggesting FoTv may have facilitated a well-regulated immune response supporting memory B-cell production, and leading to sustained, long-lasting immunity. However, effects of FoTv on Ab levels may not be solely B-cell mediated. In a Phase I clinical trial in women with breast cancer, a dose-dependent effect of Tv (a component of FoTv) on CD8+ T-cell counts was observed^34^. This suggests that augmentative effect of Tv on vaccination may occur, at least in part, via its effect on CD8+ T-cells^57^. Further, lack of Ab decay may suggest a more gradual immune response and mitigation of vaccine-triggered overproduction of acute phase inflammatory mediators (e.g., IL-1, IL-6, TNF-α). A natural adjunct that does not compromise (and potentially increases) Ab response durability could reduce vaccine hesitancy due to lessened need for/frequency of boosters. Moreover, an adjunct that reduces vaccine-related side effects could further reduce vaccine hesitancy. Such an adjunct would be highly desirable both for future COVID-19 strains and other emerging infectious diseases for which vaccination (e.g., mRNA vaccines) will likely constitute an important part of the collective strategy.

A critical problematic feature of COVID-19 has been its continuous spawning of new, mutant strains with potentially greater transmissibility or lethality than the parent strain. Such viral mutability is also likely to be characteristic of other emerging infectious diseases (e.g., H5N1). Like Ichinohe et al., we found a fungal adjunct to vaccination positively impacted Ab levels.

Ichinohe et al. additionally found that intranasal coadministration of fungal adjuvant with H5N1 vaccination not only boosted antibody production but also induced cross-protection (i.e., without requiring specific pathogen recognition) against both homologous and heterologous H5N1 strains^27^. They also found that this effect occurred because of mechanisms that were both MyD88-dependent (e.g., mediated by toll-like-receptors) and MyD88-independent (e.g., mediated by dectin-1 receptors [which bind fungal polysaccharides]). The latter finding raises the prospect that fungal mycelia may have broadly activate multiple innate immune pathways and may serve as universal immune modulators that could strengthen overall host defense systems. Our results, taken together with those of Ichinohe et al., suggest that use of a fungal mycelial adjunct to vaccination may not only increase Ab levels, but might also be predicated upon mechanisms that could generalize to future vaccines for emerging infectious diseases and to mutant strains of these viruses as well.

Fungal mycelial products may be practical for widespread, standardized use as vaccine adjuncts/adjuvants for a number of reasons:(1) because fungi are chemically complex and biosynthesize numerous compounds, they may work synergistically to address several aspects of innate and acquired immunity; (2) they may constitute a safe, natural, sustainable, scalable, low-cost, and potentially immunomodulatory strategy to reduce side effects, while preserving (and possibly increasing) Ab responses; and (3) the technology that employs aseptic cultivation and solid-state fermentation already exists, allowing for safe, rapid, and high-volume production. It should be noted that large-scale medical use of mushroom fruit bodies may not be feasible due to impracticality of controlled cultivation, concerns about their overharvesting,^58^ and potential for contamination with algae/other microorganisms^59^. Conversely, the use of cultivated mycelia as a natural vaccine adjunct constitutes a realizable, standardizable, and scalable strategy for population-level pandemic preparedness.

This study had several limitations warranting discussion. Because of rapidly changing nature of the COVID-19 pandemic, the protocol was unable to begin recruitment until after winter 2020/2021 COVID-19 peak (after initial vaccine rollout). Earlier launch might have resulted in a larger proportion of COVID-Naive participants. Because it was not logistically feasible to determine incidence of SARS-CoV-2 infection post-vaccination, biomarkers of infection risk (RBD/Spike Ab levels) were employed instead. Moreover, because Ab levels beyond six months were not examined, the extent of FoTv’s long-term effects may not have been fully appreciated (Figure 3). Additionally, other markers (e.g., inflammatory cytokine, memory B-and T-cells) that could have better elucidated the nature, or possible broadening, of the immune response were not examined. Finally, it was not feasible to examine a variety of other dosage/timing regimens.

In conclusion, this is, to our knowledge, the first human clinical trial of a fungal-based natural product as an adjunct to human vaccination and, more specifically, the first one to employ fungi in conjunction with mRNA-based or other vaccines for COVID-19. This early phase clinical trial demonstrated the safety and feasibility and provided a preliminary estimate of efficacy (short-term side effects/Ab responses), of FoTv as an adjunct to COVID-19 vaccination. In the process, it addressed and provided possible proof of the concept that fungi may simultaneously reduce vaccine reactogenicity without compromising (and possibly while increasing) immunogenicity. COVID-19 afforded the opportunities to:(a) test the strategy of using fungi as an adjunct to vaccination, and (b) do so in the midst of a pandemic caused by a novel virus.

More research is needed to elucidate precise mechanisms of fungal immunomodulation and specific clinical applications of fungi. However, the basic approach of using fungi as vaccine adjuncts appears to be safe and feasible, can be brought to bear rapidly, and may work with both mRNA and non-mRNA vaccines as well as with vaccines for other diseases.^27^ For example, because Fo mycelium has documented antiviral activity against H5N1,^45,48^ FoTv might also serve as a candidate therapeutic for H5N1. Like SARS-CoV-2 when it first appeared, H5N1 is a virus to which the human population is largely immunologically naive, and which could present suddenly and globally. These considerations take on even greater significance given the potential harm and disruption that could be posed by emerging viruses and infectious diseases.

## METHODS

### Study Design

This was a 6-month, single-site, double-blind, placebo-controlled, randomized clinical trial. Participants were recruited from and consented at COVID-19 community vaccination sites in San Diego, California. Ethics approval was obtained from the University of California San Diego. The trial protocol can be accessed at ClinicalTrials.gov, Identifier:NCT04939415^52^.

### Participants

Participants consisted of 90 eligible individuals receiving COVID-19 vaccination in the San Diego area who provided written consent. Originally, a planned sample size of 66 was estimated based on the number needed to assess safety and feasibility. However, with additional funding, sample size was increased to 90 to permit assessment of efficacy outcomes and to better equalize the number of participants in each group (see Procedure). Computations were performed using the RMASS program by Hedeker^60^ for a model with one between and one within factor (including up to 20% dropout rate) to provide minimum power of 80%. Sex was self-reported in response to an open-ended query.

### Eligibility Criteria

<u>Inclusion</u>: (1) Scheduled to receive COVID-19 vaccination/booster; and (2) age≥18. <u>Exclusion</u>: (1) Currently using investigational agents to prevent/treat COVID-19 or immunosuppressive medication; (2) known renal/hepatic disease; (3) pregnant/lactating women; and (4) prisoners.

### Randomization and Masking

Participants were randomly assigned to receive FoTv or Placebo on the day of COVID-19 vaccination (day 1) (Figure 1). Funding constraints dictated initial randomization to FoTv vs. Placebo groups in a 2:1 ratio, later adjusted to 1:1 after securing additional funding to facilitate equalizing the number of participants in each group and permit increased power for analyses of efficacy outcomes. Prior to recruitment, the statistician (S.G.) created a randomization table using block sizes of 4 and 6 (SPSS software). Only the statistician was unblinded to participant randomization. FoTv and Placebo capsules were visually identical and were distributed in unlabeled containers.

### Procedures

Short-term vaccine side effects were collected on days 1-5 (for side effect efficacy analyses) because side effects resolve in a few days^5^. To assess safety, blood from baseline and day 14 was used to measure renal/hepatic function, and adverse events were examined in this timeframe. To assess feasibility, medication adherence (Days 1-4) and completion rate at Day 14 were examined. To assess efficacy, SARS CoV-2 IgG Ab levels were obtained at the following timepoints:(1) single-dose vaccine (Johnson & Johnson) blood was collected on days 1, 14, and 28, and at 6-months post-vaccination; (2) two-dose vaccine (Pfizer-BioNTech/Moderna), on days 1, 14, and 42 (two weeks after second dose), and at 6-months post-vaccination; and (3) vaccine booster (Pfizer-BioNTech/Moderna), on days 1, 3, and 14, and at 6-months. The 6-month timepoint was chosen because, even with a 3-5 month half-life from peak titer, most individuals would still have measurable increases in Ab titer as a result of vaccination given at study onset^61^. FoTv consisted of freeze-dried FoTv mycelium fermented on a brown rice substrate, and inert Placebo of freeze-dried cooked organic brown rice. FoTv and Placebo were developed, encapsulated to a cGMP standard, and provided by Fungi Perfecti, LLC (Supplemental Materials). Capsules were packaged and appeared identical to untrained study participants.

FoTv/Placebo dosage was eight capsules (500 mg/capsule) TID for four consecutive days (starting day of first vaccination/booster). It was based on a prior study of effects of Tv on immune response^34^ and was the regimen employed in a clinical trial of FoTv in patients with active COVID-19 infection^52^. A short-term, 4-day, dosing regimen was selected to:(1) minimize subject burden and maximize adherence; and (2) permit delivery of FoTv’s expected immunomodulatory effects when vaccine-expressed Ag is present and when vaccine-related, short-term side effects are greatest.

At end of enrollment, COVID Exposure Status was ascertained from baseline blood specimens. Those with detectable anti-SARS-CoV-2 Abs (from prior COVID-19 infection or vaccination) were classified as “COVID-Exposed” and those with undetectable anti-SARS-CoV-2 Abs (mean concentration of nucleocapsid [N] protein <3 BAU [binding antibody units]/mL and mean concentration of RBD or Spike proteins <4 BAU) as “COVID-Naive”.

### Outcomes

<u>Safety:</u>(1) Adverse event (hospitalizations; ER or urgent care visits) frequency across days 1-14; and (2) serum renal/hepatic function markers on days 1 and 14. Participants were classified as having normal vs. abnormal renal function using estimated glomerular filtration rate^62^ (eGFR) and normal vs. abnormal hepatic function based on serum aspartate aminotransferase, alanine transaminase, and alkaline phosphatase. Cutoff scores for normal vs. abnormal classifications appear in Supplemental Table 1.

<u>Feasibility:</u>(1) Completion rate at Day 14 (end of safety assessment period); and (2) Treatment Adherence= (capsules taken/assigned) * 100%.

<u>Efficacy:</u>(1) Duration and severity of COVID-19 vaccine/booster side effects; and (2) Ab levels. Participants rated side effects as 0 (none or absent) to 4 (very severe) daily for Days 1-5. All CDC vaccine-associated side effects^5^ were included: Feeling feverish, low-grade afternoon fever, alternating fever or chills, chills, fatigue, muscle aches, nausea, headaches, redness, injection site swelling, and injection site pain (Supplemental Table 2). Side effect summary outcome measures were computed each day by:(1) counting total side effects present (Side effect Count); and (2) summing total side effect severities (Side Effect Severity). Abs included serum SARS-CoV-2 RBD and Spike protein levels at Days 1, 3, 14, 28/42, and 6 months.

### Data Management and Statistical Analysis

Data was managed, and statistical analyses were performed, by Krupp Center for Integrative Research (KCIR) biostatistics and data management core, using the KCIR Data System. Each participant was assigned a sequential participant number. Baseline group demographic and clinical feature comparability was tested using Analysis of Variance and Chi-square analyses. Descriptive analyses assessed data distribution, normality, and homogeneity. Missing data and group dropout rate were examined for randomness.^63^ Data were analyzed in SPSS (v.28) using two-tailed statistical tests (differences considered statistically significant for p-values <0.05).

Safety analyses were based on percentage (95% CI) of participants transitioning from normal renal/hepatic function on Day 1 to abnormal on Day 14. Feasibility used mean and 95% CIs to ascertain whether 80% completion/adherence was attained. Efficacy results employed linear mixed-effects (LME) analyses. The first analysis compared side effect levels between two grouping factors, (1) Treatment Group (FoTv vs. Placebo) and (2) COVID Exposure Status (COVID-Naive vs. COVID-Exposed), across five time points (Days 1-5). The second analysis compared Abs between the same grouping factors, across four time points (Days 3, 14, 28/42, and 6 months). To explore Ab response durability, Ab values at time-of-peak response (based on Placebo group), were compared with those at end-of-study (6 months), within each COVID Exposure Status stratum. Exploration of interaction effects was confined to analyses involving Treatment Group. Because Ab levels from Day 1 were used to create COVID-Exposure Status strata, longitudinal analyses excluded Day 1 data.

## ARTICLE INFORMATION

### Disclaimer

None.

### Data Availability

Data are available upon request.

### Additional Contributions

We would like to acknowledge the advocacy and encouragement of Lee Stein, Esq., without which the MACH-19 studies would not have been possible. We would also like to acknowledge the pioneering work and contributions of Paul Stamets, which were integral to the development of the MACH-19 studies.

### Contributors

GS, SW, and SG contributed to conceptualization; SG and TS contributed to data curation; SG, CNS, and SW carried out the formal analysis; GS and LM contributed to funding acquisition; TS, PS, and SG contributed to investigation; GS, SG, SW, AS, and LK contributed to methodology; TS, PS, GS, and LM contributed project administration; TS, SG, PS, and RD contributed resources; SG contributed software; GS, TS, LM, and DS supervised the work; SG and CNS validated the data; CNS and GS contributed to visualization; GS, CNS, SW, ZB, and CB wrote the original draft; and all authors contributed to reviewing and editing the final draft.

## Data Availability

All data produced in the present study are available upon reasonable request to the corresponding author.

