## Supplemental Materials for "Polypore Mushroom Mycelia as an Adjunct to COVID-19 Vaccination: A Randomized Clinical Trial"

**FoTv Formulation**

***Background***

Early in the COVID-19 pandemic, the authors were interested in the potential of mycelium-based products to ameliorate COVID-19 clinical symptoms; later, interest grew in the potential of mushroom mycelium and fermented substrates to mitigate adverse effects and enhance efficacy of mRNA-based SARS-CoV2 vaccines. Two polypore mushroom species, Agarikon (*Fomitopsis officinalis*, Fo) and Turkey Tail (*Trametes versicolor*, Tv), were selected based on preclinical and clinical immunological and antiviral data. The FoTv formula consists of equal components of Fo and Tv, described below.

***Materials & Methods***

Fo and Tv mycelia were independently cultivated through solid state fermentation on an organic brown rice substrate. Each myceliated fermented substrate ingredient was frozen, dried, and milled into a powder before being combined evenly and encapsulated in 500-mg pullulan capsules to produce the FoTv formulation, which was granted Investigational New Drug (IND) status by the Food and Drug Administration (FDA). An unfermented brown rice substrate, processed in the same manner, was employed as the placebo in this clinical trial.

| Supplemental Table 1. Cut off scores for identifying normal and abnormal values for liver and renal function | | |
| --- | --- | --- |
|  | Normal | Abnormal |
| Liver Function |  |  |
| Aspartate Aminotransferase (AST) | ≤ 40 U/L | > 40 U/L |
| Alanine Transaminase (ALT) | ≤ 41 U/L | > 41 U/L |
| Alkaline Phosphatase (ALP) | ≤ 129 U/L | > 129 U/L |
| Renal Function |  |  |
| Adjusted glomerular filtration rate (Adj. eGFR) | > 60 mL/min | ≤ 60 mL/min |
| *Note*. Adj. eGFR = 142 X minimum (standardized serum creatinine/K OR 1)^α^ X maximum(standardized serum creatinine/K OR 1)^-1.200^ X 0.9938^Age^ X 1.012 [if female]. Serum creatinine = mg/dL. K = 0.7 (females) or 0.9 (males). α = -0.241 (females) or -0.302 (males). Equation based on National Kidney Foundation guidelines (https://www.kidney.org/ckd-epi-creatinine-equation-2021-0). | | |

| Supplemental Table 2. Treatment Group by Day Interaction Effects for Vaccine Side effects and Other Side effects | | | | | | |
| --- | --- | --- | --- | --- | --- | --- |
|  |  | COVID-Naive Group | |  | COVID-Exposed Group | |
| Side effect |  | F value | P value |  | F value | P value |
| Vaccination Side effects | | | | | | |
| Feeling feverish |  | 2.055 | 0.092 |  | 0.972 | 0.424 |
| Low fever in the afternoon |  | 0.897 | 0.468 |  | 0.651 | 0.627 |
| Alternating fever and chills |  | 1.063 | 0.378 |  | 0.842 | 0.500 |
| Chills |  | 1.845 | 0.124 |  | 0.249 | 0.910 |
| Fatigue |  | 1.767 | 0.141 |  | 1.287 | 0.276 |
| Muscle aches |  | 1.132 | 0.346 |  | 0.852 | 0.494 |
| Nausea |  | 2.918 | 0.025 |  | 1.190 | 0.316 |
| Headaches |  | 2.009 | 0.099 |  | 0.645 | 0.631 |
| Redness/swelling at injection site |  | 0.105 | 0.981 |  | 0.473 | 0.755 |
| Pain at injection site |  | 0.932 | 0.449 |  | 0.475 | 0.754 |
| Side effect Count |  | 2.956 | 0.023 |  | 0.436 | 0.782 |
| Side effect Severity |  | 1.780 | 0.138 |  | 0.551 | 0.699 |
| Other Side effects | | | | | | |
| Belly bloat |  | 0.893 | 0.471 |  | 0.464 | 0.762 |
| Bitter taste |  | 0.489 | 0.744 |  | 0.795 | 0.529 |
| Chest fullness |  | 1.555 | 0.192 |  | 1.210 | 0.308 |
| Cold limbs |  | 2.270 | 0.065 |  | 1.916 | 0.109 |
| Diarrhea |  | 2.458 | 0.050 |  | 0.546 | 0.702 |
| Dizziness |  | 2.709 | 0.034 |  | 1.121 | 0.348 |
| Dry cough |  | 1.248 | 0.295 |  | 0.968 | 0.426 |
| Excess sweating |  | 0.458 | 0.766 |  | 1.390 | 0.239 |
| Excessive thirst |  | 1.283 | 0.281 |  | 0.435 | 0.783 |
| Heartbeat |  | 1.177 | 0.324 |  | 1.686 | 0.155 |
| Insomnia |  | 0.863 | 0.489 |  | 1.253 | 0.290 |
| Loose stools |  | 0.492 | 0.741 |  | 1.543 | 0.191 |
| Loss appetite |  | 0.365 | 0.833 |  | 0.811 | 0.519 |
| Nervousness |  | 1.199 | 0.316 |  | 0.375 | 0.826 |
| Restlessness |  | 3.907 | 0.005 |  | 1.055 | 0.380 |
| Runny nose |  | 0.649 | 0.629 |  | 1.417 | 0.230 |
| Shortness breath |  | 2.162 | 0.078 |  | 1.047 | 0.384 |
| Shortness breath exertion |  | 0.000 | 1.000 |  | 0.403 | 0.806 |
| Skin rashes |  | 1.616 | 0.175 |  | 1.809 | 0.128 |
| Sore throat |  | 0.430 | 0.786 |  | 1.630 | 0.168 |
| Stuffy nose |  | 1.141 | 0.342 |  | 0.875 | 0.480 |
| Swelling |  | 1.790 | 0.137 |  | 0.400 | 0.809 |
| *Note*. The degrees of freedom for the COVID-Naive comparisons were 4, 106 and for the COVID-Exposed comparisons were 4, 202. | | | | | | |

*Note.* Means and SDs are reported. Antibodies are in BAU (binding antibody units)/mL. Day 3 measure of Ab was obtained only for those who received a vaccine booster (and who were previously exposed via prior vaccination).

| Supplemental Table 3. Descriptive Statistics for Side effect and Antibody Measures | | | | | | |
| --- | --- | --- | --- | --- | --- | --- |
|  |  | FoTv | |  | Placebo | |
|  |  | COVID  Naive  (n=19) | COVID  Exposed  (n=30) |  | COVID  Naive  (n=11) | COVID  Exposed  (n=25) |
| Side effects | | | | | | |
| Count |  |  |  |  |  |  |
| Day 1 |  | 0.24 (0.54) | 0.65 (1.70) |  | 0.45 (0.52) | 0.48 (0.75) |
| Day 2 |  | 1.38 (2.18) | 2.87 (2.35) |  | 1.27 (2.69) | 3.19 (2.83) |
| Day 3 |  | 2.10 (2.19) | 3.13 (2.69) |  | 4.55 (3.21) | 3.26 (2.63) |
| Day 4 |  | 1.00 (1.64) | 1.45 (2.16) |  | 1.91 (1.70) | 2.04 (2.53) |
| Day 5 |  | 0.10 (0.30) | 1.00 (1.61) |  | 0.64 (0.67) | 0.85 (1.51) |
| Severity |  |  |  |  |  |  |
| Day 1 |  | 0.24 (0.54) | 1.06 (3.13) |  | 0.45 (0.52) | 0.63 (1.11) |
| Day 2 |  | 3.06 (5.27) | 5.58 (4.49) |  | 4.00 (7.13) | 6.78 (6.80) |
| Day 3 |  | 3.52 (5.11) | 6.00 (7.21) |  | 8.36 (8.88) | 5.41 (5.29) |
| Day 4 |  | 1.24 (2.28) | 2.26 (4.34) |  | 2.55 (2.84) | 3.30 (5.36) |
| Day 5 |  | 0.10 (0.30) | 1.35 (2.47) |  | 0.91 (1.22) | 1.15 (2.54) |
| Antibody | | | | | | |
| RBD |  |  |  |  |  |  |
| Day 1 |  | 1.3 (1.9) | 355.0 (358.1) |  | 1.4 (2.5) | 206.2 (188.9) |
| Day 3 |  |  | 541.9 (456.8) |  |  | 289.8 (217.4) |
| Day 14 |  | 700.6 (2530.5) | 10116.1 (6190.4) |  | 168.2 (162.0) | 10053.0 (4834.7) |
| Day 29/48 |  | 3953.5 (3460.1) | 9675.0 (5860.4) |  | 5129.0 (5704.4) | 10469.3 (3844.3) |
| 6 months |  | 6178.4 (8273.7) | 3209.5 (3720.2) |  | 3359.3 (5108) | 3398.5 (4093.5) |
| Spike |  |  |  |  |  |  |
| Day 1 |  | 0.5 (0.4) | 247.6 (258.5) |  | 0.8 (1.0) | 149.4 (120.0) |
| Day 3 |  |  | 380.7 (307.5) |  |  | 205.4 (143.3) |
| Day 14 |  | 392.5 (1047.7) | 4488.1 (1542.1) |  | 224.9 (242.1) | 4877.9 (1549.0) |
| Day 29/48 |  | 1958.7 (1542.2) | 3981.3 (1366.7) |  | 2383.3 (2275.4) | 4719.5 (1192.6) |
| 6 months |  | 2107.2 (2696.4) | 1544.4 (1341.9) |  | 1400.1 (1673.3) | 1546.5 (1347.9) |
